# Implementing the National Alzheimer’s Coordinating Center Uniform Data Set (v3) within the Diabetes Prevention Program Outcomes Study

**DOI:** 10.64898/2026.07.17.26357765

**Authors:** Lindsay K Doherty, Isabella Dechiario, Hanna Sherif, Anna Bowers, Dianilka Martinez, Danurys L Sanchez, Gerardo J Febres, Owen Carmichael, Vallabh Shah, Neelesh K Nadkarni, Terry E Goldberg, James Noble, Jose A Luchsinger, Marinella Temprosa, DPPOS Research Group

**Affiliations:** Biostatistics Center and Department of Biostatistics and Bioinformatics, Milken Institute of Public Health, George Washington University, Bethesda, MD; Departments of Medicine and Epidemiology, Columbia University Irving Medical Center, New York, NY; Department of Neurology, Columbia University Irving Medical Center, New York, NY; Pennington Research, Baton Rouge, LA; Department of Int Medicine, University of New Mexico, HSC, Albuquerque, NM; Department of Medicine (Division of Geriatric Medicine), and Neurology, University of Pittsburgh, Pittsburgh, PA; Department of Psychiatry, Columbia University Irving Medical Center, New York, NY; Taub Institute for Research on Alzheimer Disease and the Aging Brain, and G.H. Sergievsky Center, Columbia University Irving Medical Center, New York, NY

**Keywords:** Epidemiological methods, Longitudinal cohort studies, Observational studies, Neuropsychological assessment, Population based studies

## Abstract

**INTRODUCTION:** The Diabetes Prevention Program (DPP) was a randomized clinical trial designed to prevent type 2 diabetes (T2D) in adults with prediabetes. The DPP Outcomes Study (DPPOS) is the 30-year follow-up of this cohort, focusing on T2D, prediabetes, and related complications. Cognitive assessments began in 2009 and expanded in 2022 to examine cognitive impairment, including Alzheimer’s disease (AD) and AD related dementias (ADRD), in the surviving cohort. To support these aims, the National Alzheimer’s Coordinating Center Uniform Data Set version 3 (NACC-UDSv3), the standardized framework used by Alzheimer’s Disease Research Centers, was implemented in DPPOS in 2022 to enable data sharing with NACC. These forms were complemented by cognitive tests administered in DPPOS. We aimed to integrate the NACC-UDSv3 into the existing longitudinal DPPOS framework while maintaining fidelity to its structure and developing automated reports to streamline cognitive outcomes adjudication.

**METHODS:** Items from the 16 NACC-UDSv3 data forms were compared with those already collected within DPPOS to integrate overlapping similar items, add missing NACC-UDSv3 items, and create a dataset harmonized with NACC-UDSv3. Forms were adapted for electronic data capture (EDC) using the MIDAS (Multimodal Integrated Data Acquisition System, George Washington University). Automated reports integrated current and prior neuropsychological scores to support adjudications. In the first wave of the DPPOS-AD/ADRD study, 1561 cognitive adjudications were successfully completed using the harmonized DPPOS and NACC-UDSv3 data implemented into MIDAS.

**DISCUSSION:** The DPPOS-AD/ADRD project demonstrated that NACC-UDSv3 can be successfully integrated into a long-standing longitudinal cohort not originally designed for AD/ADRD research. The harmonization, electronic capture, and automated adjudication processes may provide a practical framework for other cohorts seeking to incorporate NACC-UDSv3 to align with national AD/ADRD research standards.

## Introduction

The National Alzheimer’s Coordinating Center (NACC) works with the Alzheimer’s Disease Research Centers (ADRC) Program funded by the National Institute on Aging (NIA), comprised of 35 centers across the United States. The NACC Uniform Data Set (NACC-UDS) is a standardized database comprised of multi-domain neurocognitive clinical data, phenotypic data, and criteria-based cognitive diagnoses in over 50,000 participants in the ADRC program [1].

We describe the implementation of the NACC-UDS version 3 (UDSv3) [2] forms into MIDAS (multi-model integrated data acquisition system), the existing electronic data capture (EDC) system of the Diabetes Prevention Program (DPP) Outcomes Study (DPPOS), in 2022 to expand the focus to Alzheimer’s disease (AD) and AD related dementias (ADRD). DPPOS is the observational follow-up of the DPP. DPP was originally a randomized clinical trial comparing metformin, lifestyle intervention, and placebo in the prevention of type 2 diabetes (T2D) in persons at risk [3]. In 2002, DPP transitioned to DPPOS, an observational study focused on the long-term effects of the DPP randomization arms on pre-diabetes and T2D complications [4]. In 2009, DPPOS began limited cognitive assessments of memory and executive function. In 2022, DPPOS expanded the focus with AD/ADRD. DPPOS intentionally implemented the UDSv3 forms with full fidelity to harmonize data with the NACC, and preserve longitudinal data accumulated over 25 year in a complex study that for over 25 years had a focus outside of AD/ADRD. Here we present the successful integration and implementation of the UDSv3 forms in DPPOS into MIDAS.

## Methods

### DPP and DPPOS study design

The DPP and DPPOS have been described previously [3–7]. The cohort initially randomized overweight/obese adults with pre-diabetes and followed for over 25 years. Since 1996, the major focus of DPP and DPPOS has been the development of T2D among persons with pre-diabetes and its complications. Since 2022, DPPOS aims to examine the determinants and the nature of cognitive impairment among participants with pre-diabetes and T2D, currently one third and two-thirds of the cohort, respectively. Diabetes is risk factor for cognitive impairment [8] and represent a large proportion of the U.S. population.

DPPOS-4 consists of two waves of assessments (2022-2024 and 2024-2026) over a five-year period across 25 clinical sites in the United States. Within each wave, participants complete a full study visit (Main Visit [MV]), where in addition to neurocognitive testing, participants undergo comprehensive assessments, including phlebotomy and biomarker measurement, anthropometric measures, physical function tests, and various health questionnaires. Participants were asked to have a study partner (e.g., a relative or friend), to serve as informant for UDSv3. The study accommodated participants with travel or health constraints with modified virtual visit in lieu of the in-person requirement.

### DPPOS Data Collection

Standardized data collection was performed by trained, certified Program Coordinators (PCs) and clinic personnel. Data acquisition protocols supported both traditional paper forms for subsequent transcription and direct-to-MIDAS entry, maintaining high standards of quality control across all DPPOS clinical sites. The following categories of forms have been traditionally collected in DPPOS and were implemented in the present phase:

- The <u>enrollment form</u> with sociodemographic information and family medical history information.
- <u>Study visit inventory forms</u> with information on medical history, prescription and non-prescription medications, and anthropometrics.
- <u>Procedure forms</u> with information on physical assessments and procedures performed at the main study visit.
- <u>Questionnaire forms</u> with data on physical activity, quality of life and depression, economic evaluation, social determinants of health, sexual function, sleep quality, subjective memory complaints, and legacy cognitive assessments (described below). Some questionnaires are self-administered.
- <u>Adjudication forms</u> are used to adjudicate events such as cancer, cardiovascular events (e.g. stroke), dementia, and cause of death. These forms are completed by physician adjudicators directly into MIDAS.

### Legacy cognitive assessments collected in DPPOS and DPPOS-AD/ADRD

DPPOS began cognitive testing in 2009-2010, approximately 12 years after DPP randomization, when participants were on average 63 ± 10 years of age

**Figure 1** depicts the schedule of the cognitive assessments throughout DPPOS. **Supplemental Table 1** describes all legacy cognitive assessments.

**Figure 1.**
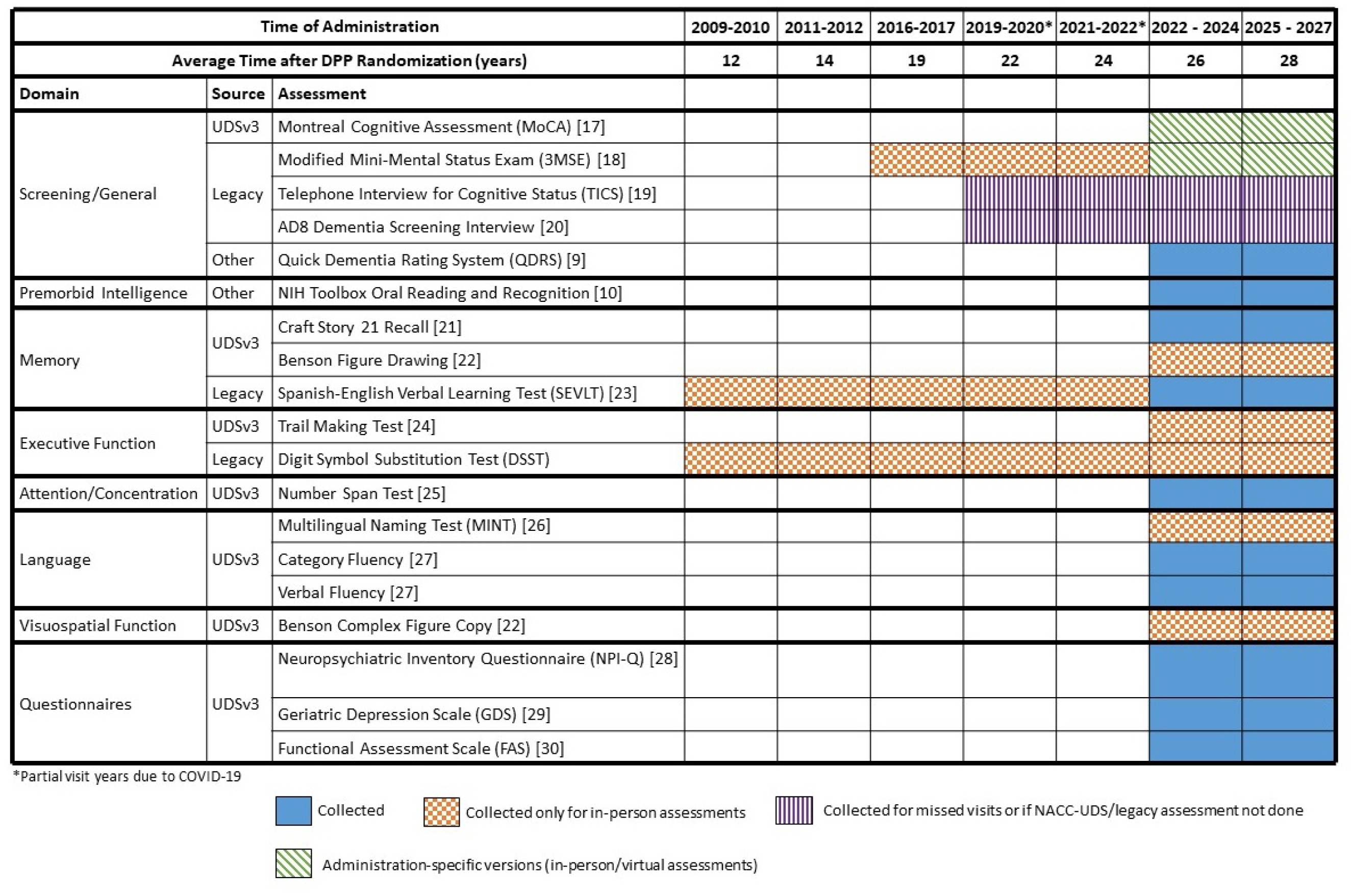
Assessments used in cognitive adjudication and timepoints of data collection.

### NACC-UDSv3 Assessments

Cognitive assessments administered in DPPOS-4 are comprised primarily of the neuropsychological battery of the UDSv3 (form C2 for in-person visits and C2T for virtual visits). The UDSv3 assessments incorporated in DPPOS are described in Figure 1 with detailed description in Supplement Table 1.

In addition to the neuropsychological battery, a neurologic exam following a script was recorded using a mobile device and read centrally by adjudicators. The adjudicators completed the neurologic exam report in the UDSv3 forms. A companion paper describes the implementation of the neurologic exam system (ref here).

### Complementary assessments not included in the UDSv3

Complementary assessments not included in the UDSv3 used for cognitive adjudications include the Quick Dementia Rating System (QDRS) [9], completed by both the participant and study partner (if available); and the NIH Toolbox Oral Reading Recognition Test [10] (administered only during in-person assessments). **Supplemental Table 1** lists descriptions of these complimentary assessments.

### Integrating NACC-UDS data forms into existing longitudinal DPPOS data collection

Existing DPPOS forms with overlapping data elements with UDSV3 were re-purposed to include required UDSv3. UDSv3 forms that did not overlap with DPPOS forms were implemented in their original format. Items from these UDSv3 forms were compared with those from the forms already collected within DPPOS to harmonize questions and maintain full fidelity to both the UDSv3 and longitudinal data collection in DPPOS. For similar but incongruent items, answer options to the DPPOS form questions were expanded to ensure that any answer marked on the DPPOS-4 form could be mapped to the corresponding UDSv3 item. UDSv3 items not already collected were added to the existing DPPOS forms. **Supplementary Table 2** lists the UDSv3 data forms and their incorporation into DPPOS-AD/ADRD.

### Overview of Cognitive Syndromes adjudicated in DPPOS-AD/ADRD

The cognitive syndrome ascertainment follows the 2011 NIA/Alzheimer’s Association (AA) recommendations for mild cognitive impairment (MCI) [11] and dementia [12] without biomarkers using the nomenclature in the UDSv3 forms. Additional standardized criteria are also applied to other common neurological disorders including Parkinson’s disease (PD) and stroke, as well as Dementia with Lewy Bodies (DLB) and frontotemporal dementia (FTD).

These syndromes are defined as follows:

- Normal cognition, defined by global clinical dementia rating (CDR) = 0 and normal cognitive testing.
- Dementia, defined by global CDR of 1 or higher (significant functional impairment) and impairment in one or more cognitive domains. Dementias are classified as: Amnestic multidomain dementia syndrome (AMDS); Posterior cortical atrophy (PCA) syndrome, following the consensus classification criteria [13]; Primary progressive aphasia (PPA), following criteria from Gorno-Tempini and colleagues [14]; Behavioral variant frontotemporal dementia (bvFTD), following revised diagnostic criteria [15]; Dementia with Lewy bodies (DLB), following the criteria of the fourth DLB consortium [16]; Non-amnestic multidomain dementia (NAMD) (not PCA, PPA, bvFTD, or DLB).
- Mild cognitive impairment (MCI), defined by a CDR = 0.5, presence of cognitive concerns, objective impairment on testing, but preserved independence of functional abilities. MCI is classified as: Amnestic MCI, single domain (only memory affected); Amnestic MCI, multi-domain (memory and another domain affected; Non-amnestic MCI, single domain (only one non-memory domain affected); Non-amnestic MCI, multi domain (> 1 non-memory domain affected).
- Cognitive impairment – not MCI. Participants who have CDR = 0.5 (considered as not normal cognition) and do not meet criteria for MCI, were defined as cognitive impairment (CI) – not MCI.

### Adjudication process and completion of physician forms of the UDSv3

Cognitive adjudication was performed by a centralized team comprising two managers, a coordinator, and five adjudicators. This group worked in close coordination with the George Washington University (GWU) Data Core, which provided the primary infrastructure for data acquisition, management, and adjudication. All study instruments and data capture workflows were hosted within MIDAS, a proprietary, custom-developed EDC system. The adjudication managers and coordinator, based at the DPPOS Cognitive Assessment and Adjudication Core at Columbia University Irving Medical Center (CUIMC), are responsible for assignment of cases to adjudicators, as well as summary and close-out of completed adjudications. All adjudicators had extensive experience with UDSv3 forms in and included two ADRC clinical core leaders. Adjudicators review assigned cases and use the EDC to directly enter the UDSv3 form used for diagnosis of cognitive syndromes (form NR1). The flow of the cognitive adjudication process is depicted in **Figure 2** and described below.

**Figure 2:**
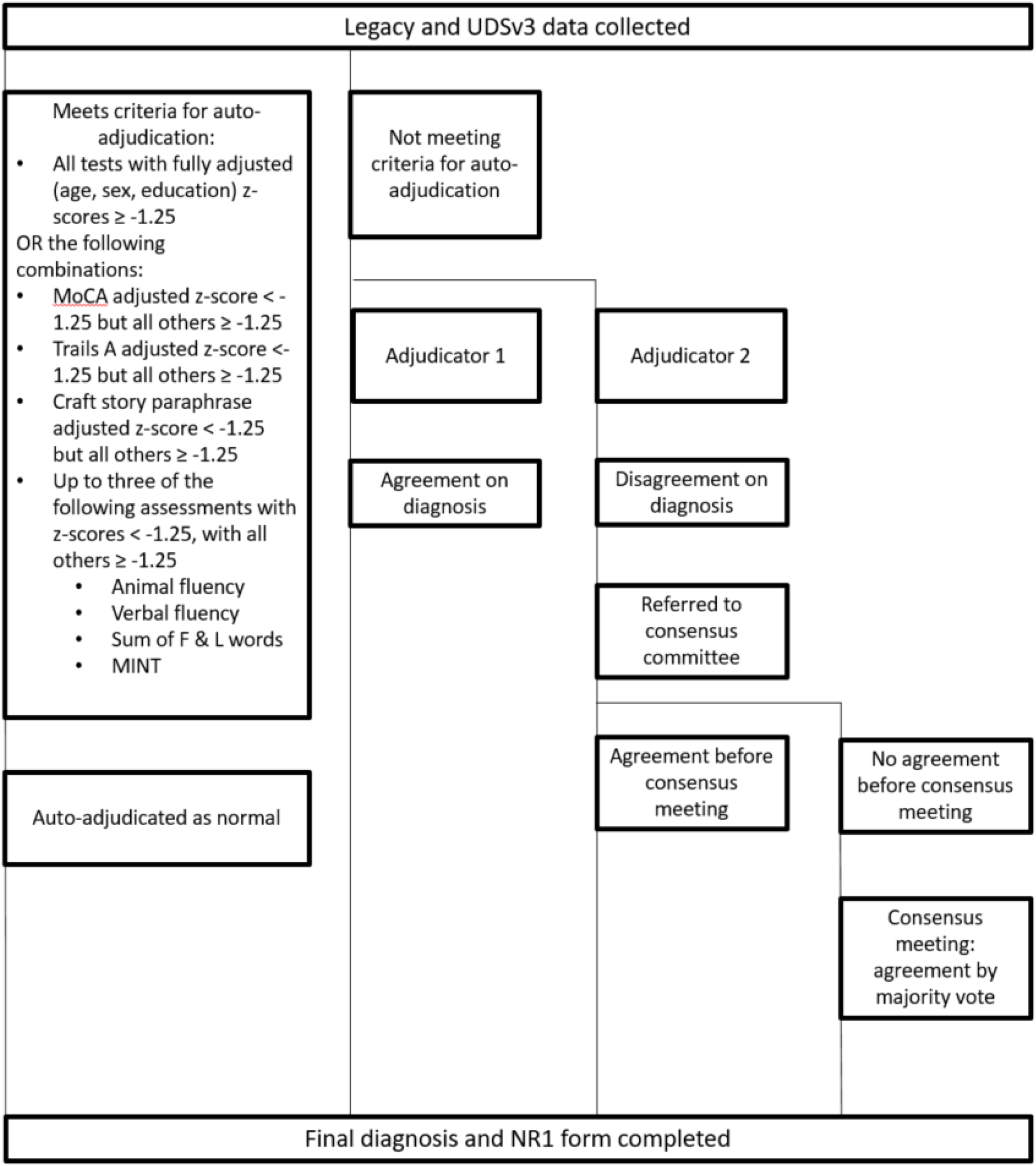
Adjudication workflow.

### Auto-adjudication

To enhance operational efficiency, an automated adjudication protocol was implemented for all assessments meeting predefined normative criteria. These cases were electronically classified as ‘normal’ and directly integrated into the EDC system. The specific algorithmic thresholds utilized for auto-adjudication are detailed in Figure 2.

### Approach to assessing performance in cognitive domains used for adjudication

The assessment encompasses six standard cognitive domains: memory, orientation, executive function, language, visuospatial function, and attention/concentration Domain-specific performance is evaluated through a standardized review process that contextualizes results within the framework of any documented visual, auditory, or physical impairments. For legacy tests, we evaluate trends in testing (stability vs. decline). For UDSv3 tests, we evaluate percentiles of performance, primarily adjusted for age, sex, and education, but we can evaluate how non-adjustment, or single adjustments, affect or bias the percentiles. In general, percentiles < 9 are considered in the impaired range. Consistency between legacy tests and the UDSv3 tests and consistency of performance in different tests within domains are also used to judge whether a domain demonstrates impairment. **Figure 1** outlines the assessments used to assess each domain.

### Cognitive Adjudication workflow

**Figure 2** describes the workflow of cognitive adjudication procedures. Participant assessments not meeting the criteria for auto-adjudication are assigned for review to two initial adjudicators. A cognitive summary report (**Figure 3**) is generated for each participant, including assessment scores, questionnaires, and longitudinal cognitive scores from UDSv3 and legacy exams, accompanied by normative data. The summary form also includes the results of informant questionnaires. Adjudicators also have access to medical history data and results from the neurological examination, not included in the summary form. Each assigned adjudicator independently completes the electronic NR1 adjudication form (**Appendix Figure 1**). After each of the two assigned adjudicators has completed the NR1 form, a consensus summary report (**Figure 4**) is generated which compares adjudicators’ classifications and answers to the NR1 form questions. A third and final NR1 is generated, pre-populated with concordant form answers between the primary and secondary adjudicator. Any form questions with discrepancies between adjudicators are left blank, to be completed during a final adjudication review. If there is agreement on diagnoses, differences in other parts of the final NR1 are resolved offline by the assigned adjudicators assisted by a manager.

**Figure 3:**
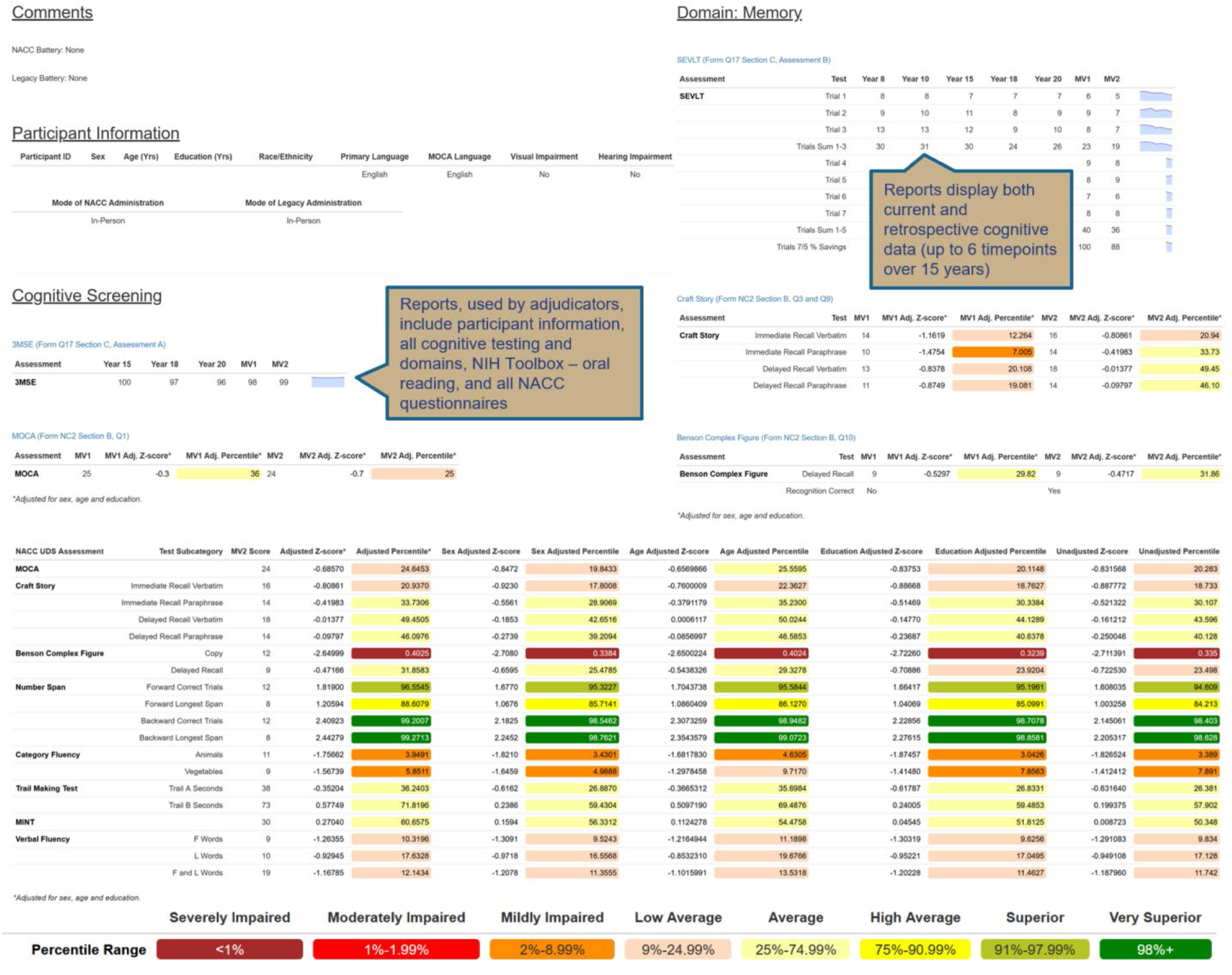
Cognitive summary report example.

**Figure 4:**
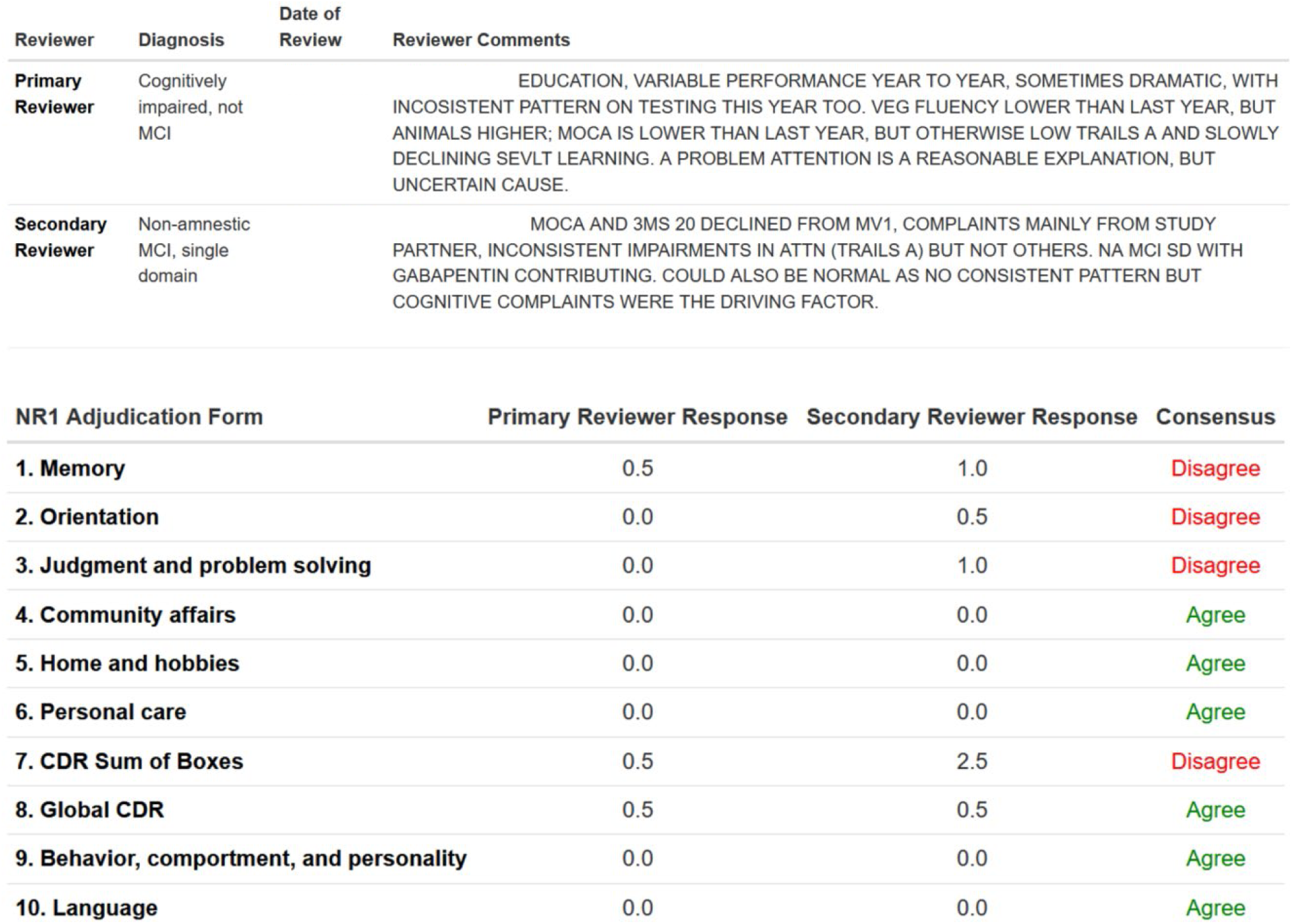
Consensus report comparing answers between adjudicators – diagnosis, rationale, answers to CDR items.

If there is disagreement on diagnoses, the cases are assigned for adjudication by the 5-member adjudication committee. The adjudication manager sends a report by email to the committee listing case IDs, adjudicators assigned, and diagnosis with comments by adjudicators. Comments include consideration of alternative diagnosis, which may lead to a change in adjudication and agreement before reaching the adjudication committee meeting. If no such agreement is achieved, cases are reviewed by the 5-member adjudication committee in one-hour remote consensus meetings. In these meetings adjudicators review the consensus summary reports, the cognitive summary reports, and a final diagnosis is decided by majority vote. The final NR1 form is completed with the information agreed to in the consensus meeting.

### Quality Control (QC) and Data Validation

All NACC-UDS assessments are double scored by PCs, either with another PC, or by the original PC at least 24 hours after the first scoring. A random sample of 10% of participants at each clinic are chosen for QC checks, in which the PC submits the paper forms from the assessments and scoring is checked by the Cognitive Assessment and Adjudication Core. Any errors are discussed with the PC, who makes corrections to scoring in the EDC.

Within MIDAS, the DPPOS forms have an extensive data validation process including skip pattern rules to ensure required fields are answered. A keyer is alerted if they attempt to skip a field that is expected or, conversely, if the keyer attempts to answer a field that should be skipped. Additionally, the coordinating center implements inter-field checks to further ensure integrity between fields. If a form keyer attempts to submit a form which violates any skip pattern rules or inter-field checks, a pop-up window shows all fields needing attention. Validation reports are also run weekly to account for any overridden skip patterns or inter-field checks, and any other more complex rules that cannot be specified within MIDAS.

### Completed Adjudications in the first wave of DPPOS-4

Of 1683 consenting participants, 1561 completing cognitive assessments were adjudicated between 2022-2024(**Table 1**). A total of 1294 (83%) participants completed in-person visits, 243 (16%) completed remote visits, and 24 (2%) completed partially in-person and partially remote visits (hybrid), and 291 (19%) were auto-adjudicated. Of the remaining 1270 participants reviewed by adjudicators, 1233 (97%) gave sufficient data to receive a diagnosis. There was a first pass agreement between the two adjudications in 673 (53%), and a second pass agreement after review of minor disagreements in 264 (21%). Finally, 333 (26%) cases went to full adjudication committee.

**Table 1.**
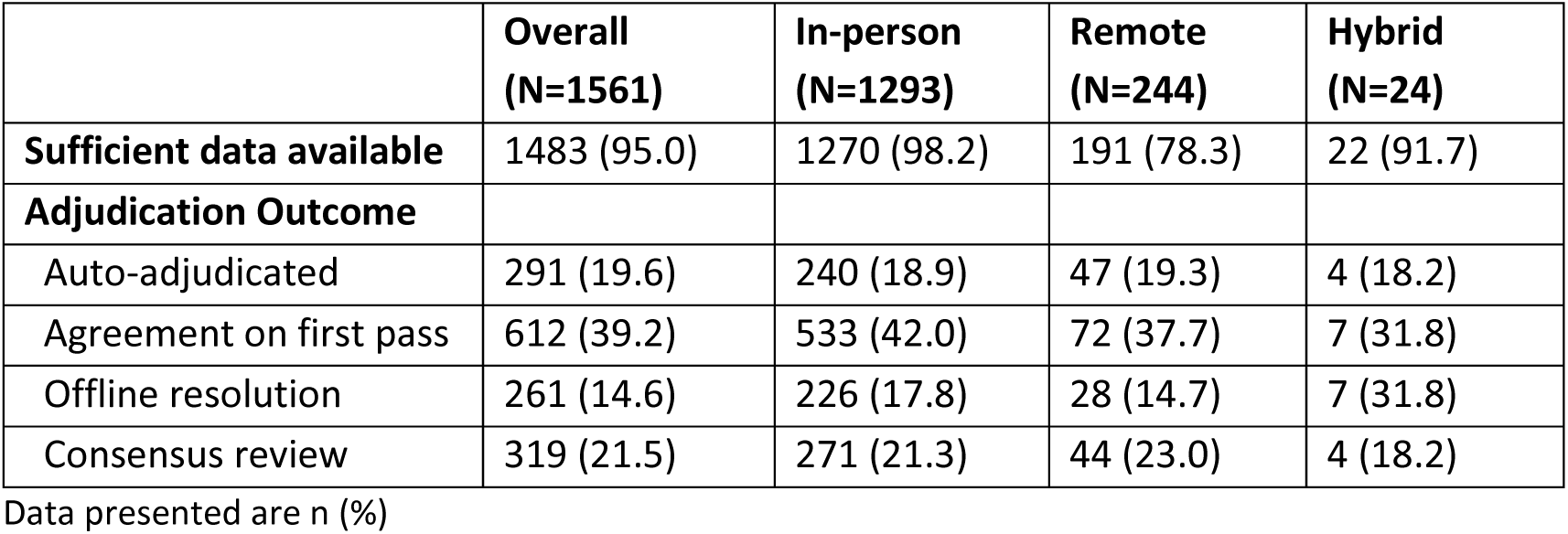
Description of adjudicated cases (N=1561)

## Discussion

We successfully implemented the UDSv3 forms with full fidelity integrating new UDSv3 elements, complementary questionnaires, legacy cognitive tests, and medical history and medication data available from DPPOS. Furthermore, we believe that this is the first time that the UDSv3 forms were integrated into a non-AD/ADRD cohort with full fidelity, wherein the datasets generated are compatible with those received by the NACC from all ADRCs in the United States. The UDSv3 data from DPPOS contributed to NACC will add valuable information on AD/ADRD in the special high-risk group of persons with T2D and pre-diabetes represented in DPPOS.

This report describes the implementation of the UDSv3 forms without AD/ADRD biomarkers. At the time of this report, collection of research brain imaging biomarkers including cerebrovascular disease on magnetic resonance imaging (MRI), measures of neurodegeneration on MRI, and amyloid positron emission tomography (PET) are underway in approximately a third of the cohort. This biomarker data will be integrated with the UDSv3 forms once available with the caveat that these biomarkers were collected for research and not clinical purposes. In addition, plasma biomarkers of amyloid burden are available in 82% of participants, which will also be integrated into the UDSv3 data.

The UDS version 4 forms were under development when DPPOS-4 started, leading to the implementation of the UDSv3 forms instead. Comparing UDSv3 and the UDSv4 D1 consensus criteria forms, concepts of normal cognitive aging, MCI, and dementia were largely similar with the main exception being the addition of behavioral and apraxia domains to the list of affected domains for persons diagnosed with MCI or dementia. The UDSv4 D1 form formalized the construct of “impaired, not-MCI”, which we implemented as “CI-not MCI”, intended to “capture those individuals with evidence of cognitive impairment or decline who do not meet formal MCI criteria.” Our criteria of CI-not MCI using the UDSv3 forms largely follows the criteria formalized in the UDSv4 forms. Subjective cognitive decline was also added to UDSv4 D1a, but this concept was not incorporated into the DPPOS adjudications as it was not part of the UDSv3 D1 form. However, DPPOS cognitive interview forms allow for a post-hoc determination of subjective cognitive decline in those participants who report various concerns but have no evidence of impairment on testing.

It is important to note that there are several limitations in our data as it compares to that collected at the ADRCs. The neuropsychological tests and other questionnaires were collected by PCs with at least a bachelor’s degree that were certified by the Cognitive Assessment and Adjudication Core. Although each ADRC differs in their processes most centers have participants undergo testing with a local research assistant who collects cognitive test data, however each ADRC trains their staff by local neuropsychologists and psychometricians rather than a central team, as done in DPPOS-AD/ADRD. The adjudication forms were completed by clinician members of the adjudication committee who did not personally encounter study participants, using summary data generated from the data collected by the PCs who did not participate in the adjudication meetings. In contrast, in the ADRC consensus meetings clinician and research team members who have personally encountered study participants personally provide their insights.

Non-AD/ADRD cohorts have the potential of providing important information that can be compared or integrated by the NACC data from ADRCs. Our report provides an example of best practices for the successful implementation of the NACC-UDS forms in a non-AD/ADRD cohort, its integration with an existing EDC system, and its integration of existing EDC forms to complement the UDSv3 forms.

## Supporting information

DPPOS Investigators Appendix

Appendix. Example of NR1 form in MIDAS

## Acknowledgement

The DPP Research Group gratefully acknowledges the commitment and dedication of the participants of the DPP and DPPOS. This project was supported by the National Institute of Diabetes and Digestive and Kidney Diseases (NIDDK) of the National Institutes of Health (NIH) under award numbers U01 DK048489, U01 DK048339, U01 DK048377, U01 DK048349, U01 DK048381, U01 DK048468, U01 DK048434, U01 DK048485, U01 DK048375, U01 DK048514, U01 DK048437, U01 DK048413, U01 DK048411, U01 DK048406, U01 DK048380, U01 DK048397, U01 DK048412, U01 DK048404, U01 DK048387, U01 DK048407, U01 DK048443, and U01 DK048400, and in part by the National Institute on Aging of the NIH under award number 5 U19 AG078558 by providing funding during DPP and DPPOS to the clinical centers and the Coordinating Center for the design and conduct of the study, and collection, management, analysis, and interpretation of the data. Funding was also provided by the National Institute of Child Health and Human Development, the National Eye Institute, the National Heart Lung and Blood Institute, the National Cancer Institute, the Office of Research on Women’s Health, the National Institute on Minority Health and Health Disparities, the Centers for Disease Control and Prevention, and the American Diabetes Association. The content is solely the responsibility of the authors and does not necessarily represent the official views of the National Institutes of Health. The Southwestern American Indian Centers were supported directly by the NIDDK, including its Intramural Research Program, and the Indian Health Service. The General Clinical Research Center Program, National Center for Research Resources, and the Department of Veterans Affairs supported data collection at many of the clinical centers. Merck KGaA provided medication for DPPOS. DPP/DPPOS have also received donated materials, equipment, or medicines for concomitant conditions from Bristol-Myers Squibb, Parke-Davis, and LifeScan Inc., Health O Meter, Hoechst Marion Roussel, Inc., Merck-Medco Managed Care, Inc., Merck and Co., Nike Sports Marketing, Slim Fast Foods Co., and Quaker Oats Co. McKesson BioServices Corp., Matthews Media Group, Inc., and the Henry M. Jackson Foundation provided support services under subcontract with the Coordinating Center. The sponsor of this study was represented on the Steering Committee and played a part in study design, how the study was done, and publication. All authors in the writing group had access to all data. The opinions expressed are those of the study group and do not necessarily reflect the views of the funding agencies. A complete list of Centers, investigators, and staff can be found in the Appendix.

List of Funding Agencies: NIDDK, IHS, NCRR, Dept. VA, NICHD, NIA, NEI, NHLBI, NCI, ORWH, NIMHHD, CDC, ADA, Merck KGaA, Bristol-Myers Squibb, Parke-Davis, Lipha, Lifescan.

## Statement of Ethics

The study was conducted according to the guidelines set out in the Declaration of Helsinki and all procedures involving research study participants. Prior to initiating the study protocol, each participant provided written informed consent and each study center obtained approval from its respective institutional review board. The trials are registered at ClinicalTrials.gov (Diabetes Prevention Program: NCT00004992; Diabetes Prevention Program Outcomes Study: NCT00038727; Diabetes Prevention Program Outcomes Study AD/ADRD: NCT05704309).

## Author Contributions

ID, HS, AB, DM, DS, GF, OC, VS, NN, TG, JN, and MT edited and reviewed the manuscript, and contributed to discussion. LD and JL wrote the manuscript and LD researched the data.

## Data Availability Statement

In accordance with the NIH Public Access Policy, we continue to provide all manuscripts to PubMed Central including this manuscript DPP/DPPOS has provided the protocols and lifestyle and medication intervention manuals to the public through its public website (https://www.dppos.org). The DPPOS abides by the NIDDK data sharing policy and implementation guidance as required by the NIH/NIDDK (https://repository.niddk.nih.gov/studies/dppos/)

## References

1. NACC-UDS, University of Washington School of Public Health. https://www.naccdata.org/. Accessed Mar 10, 2026.

2. Besser L, Kukull W, Knopman DS, et al. Version 3 of the national alzheimer’s coordinating center’s uniform data set. Alzheimer Dis Assoc Disord. 2018;32(4):351–358. doi: 10.1097/WAD.0000000000000279.

3. Knowler WC, Barrett-Connor E, Fowler SE, et al. Reduction in the incidence of type 2 diabetes with lifestyle intervention or metformin. N Engl J Med. 2002;346(6):393–403. doi: 10.1056/NEJMoa012512.

4. Diabetes Prevention Program Research Group, Knowler WC, Fowler SE, et al. 10-year follow-up of diabetes incidence and weight loss in the diabetes prevention program outcomes study. Lancet. 2009;374(9702):1677–1686. doi: 10.1016/S0140-6736(09)61457-4.

5. Diabetes Prevention Program Research Group. Long-term effects of lifestyle intervention or metformin on diabetes development and microvascular complications over 15-year follow-up: The diabetes prevention program outcomes study. Lancet Diabetes Endocrinol. 2015;3(11):866–875. doi: 10.1016/S2213-8587(15)00291-0.

6. Heckman-Stoddard BM, Crandall JP, Edelstein SL, et al. Randomized study of metformin and intensive lifestyle intervention on cancer incidence over 21 years of follow-up in the diabetes prevention program. Cancer Prev Res (Phila). 2025;18(7):401–411. doi: 10.1158/1940-6207.CAPR-23-0461.

7. Goldberg RB, Orchard TJ, Crandall JP, et al. Effects of long-term metformin and lifestyle interventions on cardiovascular events in the diabetes prevention program and its outcome study. Circulation. 2022;145(22):1632–1641. doi: 10.1161/CIRCULATIONAHA.121.056756.

8. 2025 Alzheimer’s disease facts and figures. Alzheimers Dement. 2025 Apr 29;21(4):e70235. doi: 10.1002/alz.70235. PMCID: PMC12040760.

9. Galvin JE. The quick dementia rating system (qdrs): A rapid dementia staging tool. Alzheimers Dement (Amst). 2015;1(2):249–259. doi: 10.1016/j.dadm.2015.03.003.

10. Gershon RC, Cook KF, Mungas D, et al. Language measures of the NIH toolbox cognition battery. J Int Neuropsychol Soc. 2014;20(6):642–651. doi: 10.1017/S1355617714000411.

11. Albert MS, DeKosky ST, Dickson D, et al. The diagnosis of mild cognitive impairment due to alzheimer’s disease: Recommendations from the national institute on aging-alzheimer’s association workgroups on diagnostic guidelines for alzheimer’s disease. Alzheimers Dement. 2011;7(3):270–279. doi: 10.1016/j.jalz.2011.03.008.

12. McKhann GM, Knopman DS, Chertkow H, et al. The diagnosis of dementia due to alzheimer’s disease: Recommendations from the national institute on aging-alzheimer’s association workgroups on diagnostic guidelines for alzheimer’s disease. Alzheimers Dement. 2011;7(3):263–269. doi: 10.1016/j.jalz.2011.03.005.

13. Crutch SJ, Schott JM, Rabinovici GD, Murray M, Snowden JS, et al. Alzheimer’s Association ISTAART Atypical Alzheimer’s Disease and Associated Syndromes Professional Interest Area. Consensus classification of posterior cortical atrophy. Alzheimers Dement. 2017 Aug;13(8):870–884. doi: 10.1016/j.jalz.2017.01.014. Epub 2017 Mar 2. PMID: 28259709; PMCID: PMC5788455.

14. Gorno-Tempini ML, Hillis AE, Weintraub S, Kertesz A, Mendez M, et al. Classification of primary progressive aphasia and its variants. Neurology. 2011 Mar 15;76(11):1006–14. doi: 10.1212/WNL.0b013e31821103e6. Epub 2011 Feb 16. PMID: 21325651; PMCID: PMC3059138.

15. Rascovsky K, Hodges JR, Knopman D, Mendez MF, Kramer JH, et al. Sensitivity of revised diagnostic criteria for the behavioural variant of frontotemporal dementia. Brain. 2011 Sep;134(Pt 9):2456–77. doi: 10.1093/brain/awr179. Epub 2011 Aug 2. PMID: 21810890; PMCID: PMC3170532.

16. McKeith IG, Dickson DW, Lowe J, Emre M, O’Brien JT, et al. Consortium on DLB. Diagnosis and management of dementia with Lewy bodies: third report of the DLB Consortium. Neurology. 2005 Dec 27;65(12):1863–72. doi: 10.1212/01.wnl.0000187889.17253.b1. Epub 2005 Oct 19. Erratum in: Neurology. 2005 Dec 27;65(12):1992. PMID: 16237129.

17. Nasreddine ZS, Phillips NA, Bédirian V, et al. The montreal cognitive assessment, MoCA: A brief screening tool for mild cognitive impairment. J Am Geriatr Soc. 2005;53(4):695–699. doi: 10.1111/j.1532-5415.2005.53221.x.

18. Teng, E.L., & Chui, H.C. (1987). The Modified Mini-Mental State (3MS) examination. The Journal of clinical psychiatry, 48 8, 314-8

19. Fong TG, Fearing MA, Jones RN, et al. Telephone interview for cognitive status: Creating a crosswalk with the mini-mental state examination. Alzheimers Dement. 2009;5(6):492–497. doi: 10.1016/j.jalz.2009.02.007.

20. Galvin JE, Roe CM, Coats MA, Morris JC. Patient’s rating of cognitive ability: Using the AD8, a brief informant interview, as a self-rating tool to detect dementia. Arch Neurol. 2007;64(5):725–730. doi: 10.1001/archneur.64.5.725.

21. Craft S, Newcomer J, Kanne S, et al. Memory improvement following induced hyperinsulinemia in alzheimer’s disease. Neurobiol Aging. 1996;17(1):123–130. doi: 10.1016/0197-4580(95)02002-0.

22. Possin KL, Laluz VR, Alcantar OZ, Miller BL, Kramer JH. Distinct neuroanatomical substrates and cognitive mechanisms of figure copy performance in alzheimer’s disease and behavioral variant frontotemporal dementia. Neuropsychologia. 2011;49(1):43–48. doi: 10.1016/j.neuropsychologia.2010.10.026.

23. González HM, Mungas D, Reed BR, Marshall S, Haan MN. A new verbal learning and memory test for english- and spanish-speaking older people. J Int Neuropsychol Soc. 2001;7(5):544–555. doi: 10.1017/s1355617701755026.

24. Reitan R. Trail Making Test. Manual for administration and scoring. Neuropsychological Laboratory; 1992

25. Weintraub S, Besser L, Dodge HH, et al. Version 3 of the alzheimer disease centers’ neuropsychological test battery in the uniform data set (UDS). Alzheimer Dis Assoc Disord. 2018;32(1):10–17. doi: 10.1097/WAD.0000000000000223.

26. Gollan TH, Weissberger GH, Runnqvist E, Montoya RI, Cera CM. Self-ratings of spoken language dominance: A multi-lingual naming test (MINT) and preliminary norms for young and aging spanish-english bilinguals. Biling (Camb Engl). 2012;15(3):594–615. doi: 10.1017/S1366728911000332.

27. Tombaugh TN, Kozak J, Rees L. Normative data stratified by age and education for two measures of verbal fluency: FAS and animal naming. Arch Clin Neuropsychol. 1999;14(2):167– 177.

28. Cummings JL, Mega M, Gray K, Rosenberg-Thompson S, Carusi DA, Gornbein J. The neuropsychiatric inventory: Comprehensive assessment of psychopathology in dementia. Neurology. 1994;44(12):2308–2314. doi: 10.1212/wnl.44.12.2308.

29. Yesavage JA. The use of self-rating depression scales in the elderly. In: Handbook for clinical memory assessment of older adults. Washington, DC, US: American Psychological Association; 1986:213–217. 10.1037/10057-017.

30. Pfeffer RI, Kurosaki TT, Harrah CH, Chance JM, Filos S. Measurement of functional activities in older adults in the community. Journal of gerontology. 1982;37(3):323–329. doi: 10.1093/geronj/37.3.323.

