## Supplementary figures and images for "Implementing the National Alzheimer’s Coordinating Center Uniform Data Set (v3) within the Diabetes Prevention Program Outcomes Study"

### Appendix. Example of NR1 form in MIDAS

**Appendix. Example of NR1 form in MIDAS**

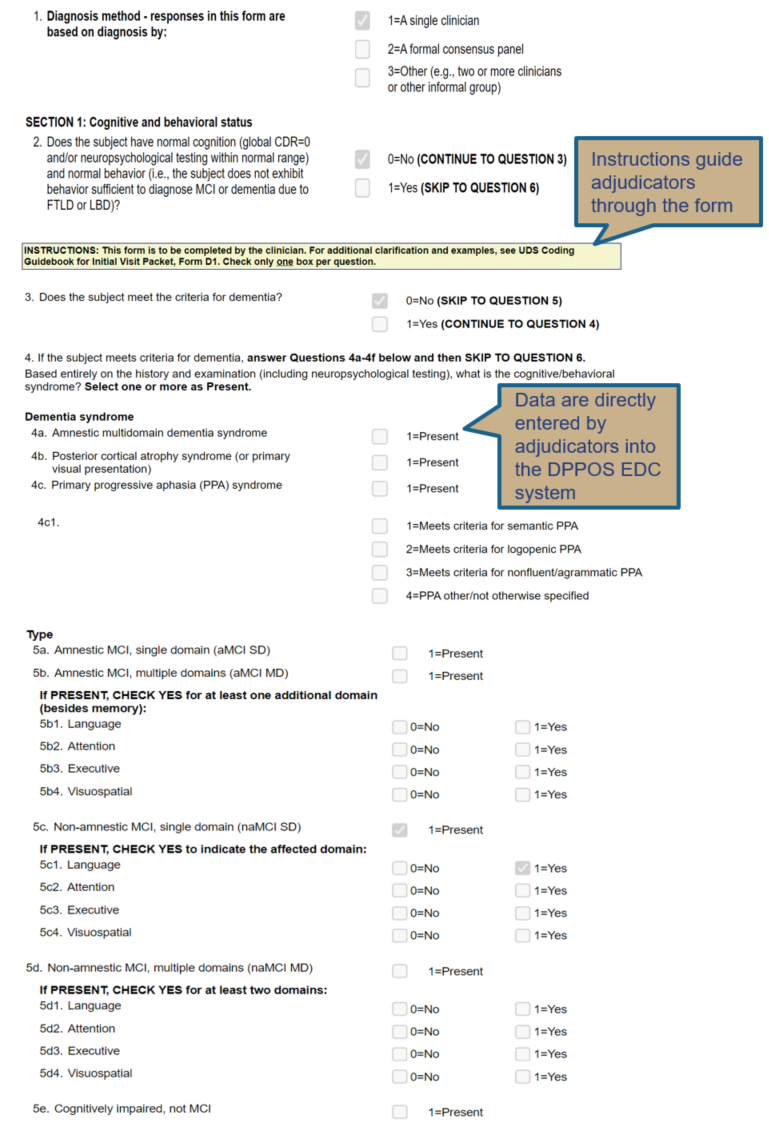
