## Supplemental Tables and Figures for "Implementing the National Alzheimer’s Coordinating Center Uniform Data Set (v3) within the Diabetes Prevention Program Outcomes Study"

Supplementary Table 1. Description of all DPPOS and DPPOS-AD/ADRD assessments

| **Domain** | **Source** | **Assessment** | **Description** |
| --- | --- | --- | --- |
| **Memory** | **NACC** | **Craft Story 21 Recall (Immediate)** | Assesses the ability to recall a short story. The examiner reads the story to the subject in a clear voice. Immediately after hearing the story, the participant is asked to retell the story from memory. The story should be read with adequate volume and clarity for the subject to understand during the presentation. No repetitions are permitted.  The participant receives two scores: (1) verbatim scoring records the number of words verbatim the participant can recite, and (2) paraphrase scoring gives alternative points for similar words used to recite the story.  Instructional text like this is probably unnecessary, at least in the body of the manuscript. |
|  |  | **Craft Story 21 Recall (Delayed)** | A test of delayed recall (episodic memory) of the story read to the subject at the beginning of the testing session, during the Craft Story 21 Recall (Immediate). The subject receives verbatim and paraphrase scoring which are collected in the same way as the Craft Story 21 Recall (Immediate) |
|  |  | **Benson Figure Drawing** | A simplified form of the Rey-Osterrieth Complex Figure. The purpose of the test is to assess a subject's visuoconstructional and visual memory functions. The participant is given a pen and a sheet with a figure and asked to copy the design as best they can. Scoring is based on a rubric for each element of the figure, and one point for whether it was drawn correctly, and another point for whether it was placed correctly. The score is the overall sum |
|  | **Legacy** | **Spanish-English Verbal Learning Test (SEVLT)** | A measure of new learning and verbal memory. The participant is asked to recall a list of 15 common words over three trials. For each trial, the participant is read the list and asked immediately afterwards to recall as many of the words as they can. In previous phases, this was performed three times with the same list of words (trials one, two, three). Next, a new distractor word list was presented (trial four) and the participant is asked to repeat each word. Immediately after the recitation of the distractor word list, the participant is asked to recall the first list again (trial five).  For DPPOS-AD/ADRD, the SEVLT was expanded to a total of seven trials. The first five trials are similar to the original first three trials, where the participant is read the 15 common words and asked immediately each time to recall as many of the words as they can (trials one through five). Next, a new distractor list is presented and the participant is asked to repeat each word (trial six). Following the distractor list, a 20-30 minute delay is administered. After the delay, the participant is asked to recall the first list again (trial seven). |
| **Executive Function** | **NACC** | **Trail Making Test** | A test of processing speed and executive function^15^. Part A consists of 25 circles numbered 1 through 25 distributed over a white sheet of 8 1/2" x 11" paper. The participant is instructed to connect the circles with a drawn line as quickly as possible in ascending numerical order. Part B also consists of 25 circles, but these circles contain either numbers (1 through 13) or letters (A through L). The participant must connect the circles while alternating between numbers and letters in an ascending order (e.g., A to 1; 1 to B; B to 2; 2 to C). The participant gets a score for the number of seconds required to complete each trail (the primary measure of interest, with a maximum allowed time of 300 seconds) and the numbers of correct lines and errors for each trail is also reported. |
|  | **Legacy** | **Digit Symbol Substitution Test (DSST)** | A measure of frontal executive abilities, psychomotor speed and sustained attention. The DSST is a test in which participants try to match numbers to symbols in 90 seconds. The total number of correct answers is reported. In this task, the participant is asked to translate numbers (1-9) to symbols using a key provided at the top of the test form |
| **Attention / Concentration** | **NACC** | **Number Span Test** | A test of working memory which taps two different working memory constructs. The first, Forward Number Span, measures the capacity for holding information very briefly for the purpose of repeating it exactly. The second, Backward Number Span, measures the ability not only to hold the information but also to manipulate the numbers and reverse the sequence. Numbers for both forward and backward span tests are presented, with sequences ranging from 2 to 9 numbers. Two trials are administered at each sequence length, each with two scores: One for the number of correct trials, and the longest span of a correct trial. |
| **Language** | **NACC** | **Multilingual Naming Test (MINT)** | A test of visual object naming designed to contain items that have similar levels of usage and familiarity across four different languages: English, Spanish, Hebrew, and Mandarin^16^. Line drawings are presented to the subject with the instruction to say the name of the object. Scores are also derived from the number the participant got correct, with or without a semantic cue (verbal hint, e.g. butterfly - "an insect"). |
|  |  | **Category Fluency** | A widely used measure of semantic memory (verbal fluency, language). The participant is asked to name as many animals as they can name in 60 seconds, and is then asked to name as many vegetables they can name in 60 seconds. The number of unique animals and vegetables are scored, for a total of two scores. |
|  |  | **Verbal Fluency** | A widely used measure of word generation that may be sensitive to dysfunction in the dominant frontal lobe. The Participant is asked to say as many words as possible that begin with the letter "F" in 60 seconds, and then as many words that begin with the letter "L" in 60 seconds. There is a score for the number of unique "F" words given, a score for the number of unique "L" words, and a score for the total number of unique "F" and "L" words. |
| **Visuospatial Function** | **NACC** | **Benson Complex Figure Copy** | A test of visuospatial abilities. Ten to 15 minutes after the Benson Complex Figure Copy trial (described below in visuospatial skills), the participant is asked to draw the picture again from memory. Then, the participant is presented a sheet with four figures on it, one of which is the picture they were originally asked to copy. The participant is asked to identify the one they were asked to copy. Two scores are given: one score is based on a rubric for each element of the figure drawn by the participant, and one point for whether it was drawn correctly, and another point for whether it was placed correctly. This score is the overall sum. The other is an indicator of whether the participant correctly identified the figure they had been instructed to draw (Yes or No). |
| **Screening/General** | **NACC** | **Montreal Cognitive Assessment (MoCA)** | A rapid screening instrument designed to help health professionals detect mild cognitive dysfunction. It assesses numerous cognitive domains: attention and concentration, executive functions, memory, language, visuoconstructional skills, conceptual thinking, calculations, and orientation |
|  | **Legacy** | **Modified Mini-Mental Status Exam (3MSE)** | A brief screening test for dementia with demonstrated reliability and validity. |
|  |  | **Telephone Interview for Cognitive Status (TICS)** | A global mental status test that can either be administered over the telephone or face-to-face, and was developed for use in situations where in-person cognitive screening is impractical or inefficient. In DPPOS-AD/ADRD, the TICS is administered only to those who do not complete both the legacy and NACC-UDSv3 cognitive assessments. Cognitive domains measured by the TICS include orientation, concentration, short-term memory, language, praxis, and mathematical skills |
|  |  | **AD8 Dementia Screening Interview** | A screening test for dementia, ideally given administered to the study partner, but can be administered to the participant if a study partner is unavailable, collects information regarding changes in cognitive status and functional abilities over the last several years. The AD8 is administered only to those who do not complete both the legacy and NACC-UDSv3 cognitive assessments. The AD8 is sensitive to detecting early cognitive changes associated with many common dementing illnesses including Alzheimer’s disease, vascular dementia, Lewy body dementia, and frontotemporal dementia. |
|  | **Other** | **Quick Dementia Rating System (QDRS)** | A 10 item questionnaire, administered to both the participant and study partner if available. Collects information on degree of cognitive and behavioral change across 10 domains: Memory and recall, orientation, decision-making and problem-solving abilities, activities outside the home, function at home and hobbies, toileting and personal hygiene, behavior and personality changes, language and communication abilities, mood, and attention and concentration |
| **Premorbid Intelligence** | **Other** | **NIH Toolbox Oral Reading and Recognition** | An assessment of reading decoding skills and crystalized abilities. Participant are asked to read aloud letters and words, pronouncing them as accurately as possible. |
| **Questionnaires** | **NACC** | **Neuropsychiatric Inventory Questionnaire (NPI-Q)** | A brief assessment of neuropsychiatric symptomatology in routine clinical practice settings, adapted from the NPI^18^. Designed to be a self-administered questionnaire by the study partner about the participants, rating the severity of 12 cognitive domains. |
|  |  | **Functional Assessment Scale (FAS)** | Also known as the Functional Activities Questionnaire (FAQ), measures instrumental activities of daily living, such as preparing balanced meals and managing personal finances. Designed to be administered by the study partner in regards to the participants. |
|  |  | **Geriatric Depression Scale (GDS)** | A scale developed as a basic screening measure for depression in older adults. |

Supplementary Table 2. NACC-UDS forms and their incorporation in to DPPOS-AD/ADRD

| **Form Number** | **UDSv3 Form Name** | **Administration in DPPOS-AD/ADRD** | **Incorporation into EDC** |
| --- | --- | --- | --- |
| A1 | Subject Demographics | Incorporated into the DPPOS-AD/ADRD Study Enrollment Forms | Paper form completed at clinic and entered into EDC |
| A4 | Subject Medications |  |  |
| A5 | Subject Health History |  |  |
| B1 | Physical |  |  |
| A2 | Co-participant Demographics | Administered in original format. |  |
| A3 | Subject Family History |  |  |
| B5 | Neuropsychiatric Inventory Questionnaire (NPI-Q) |  |  |
| B6 | Geriatric Depression Scale (GDS) |  |  |
| B7 | NACC Functional Assessment Scale (FAS) |  |  |
| D2 | Clinician-assessed Medical Conditions |  | Populated directly into EDC from adjudicated health records and participant report |
| B8 | Neurological Examination Findings |  | Entered by physicians directly into EDC |
| B4 | CDR™ Plus NACC FTLD | Combined into a single cognitive adjudication form | Entered by adjudicators directly into EDC |
| B9 | Clinician Judgment of Symptoms |  |  |
| D1 | Clinician Diagnosis |  |  |
